# Internal and external factors affecting vaccination coverage: modeling the interactions between vaccine hesitancy, accessibility, and mandates

**DOI:** 10.1101/2022.09.20.22280174

**Authors:** Kerri-Ann Anderson, Nicole Creanza

## Abstract

Society, culture, and individual motivations affect human decisions regarding their health behaviors and preventative care, and health-related perceptions and behaviors can change at the population level as cultures evolve. An increase in vaccine hesitancy, an individual mindset informed within a cultural context, has resulted in a decrease in vaccination coverage and an increase in vaccine-preventable disease (VPD) outbreaks, particularly in developed countries where vaccination rates are generally high. Understanding local vaccination cultures, which evolve through an interaction between beliefs and behaviors and are influenced by the broader cultural landscape, is critical to fostering public health. Vaccine mandates and vaccine inaccessibility are two external factors that interact with individual beliefs to affect vaccine-related behaviors. To better understand the population dynamics of vaccine hesitancy, it is important to study how these external factors could shape a population’s vaccination decisions and affect the broader health culture. Using a mathematical model of cultural evolution, we explore the effects of vaccine mandates, vaccine inaccessibility, and varying cultural selection trajectories on a population’s level of vaccine hesitancy and vaccination behavior. We show that vaccine mandates can lead to a phenomenon in which high vaccine hesitancy co-occurs with high vaccination coverage, and that high vaccine confidence can be maintained even in areas where access to vaccines is limited.

## Introduction

A comprehensive understanding of health behaviors and their potential for exacerbating or mitigating disease risk requires insight into how cultural beliefs influence these behaviors. Local vaccination cultures—the shared beliefs among individuals within a community about vaccine-preventable disease etiology, prevention, and treatment—can affect an individual’s vaccine attitudes and decisions [1,2]. The definition of “vaccine hesitancy” varies between sources, spanning from an attitude of uncertainty about vaccines to the behavior of vaccine refusal. Here we use the definition from Larson *et al*. 2022 [3]: “a state of indecision and uncertainty that precedes a decision to become (or not become) vaccinated.” In 2019, vaccine hesitancy was named one of the World Health Organization’s ten threats to global health [4] because of its link to reduced vaccination coverage and more frequent outbreaks of vaccine-preventable diseases (VPDs) worldwide. Vaccine hesitancy is a key indicator of the vaccination culture of a population, and public health studies have considered vaccine hesitancy to be influenced by multiple societal- and individual-level factors, such as the vaccination coverage of the population, the perceived risk of vaccine-preventable diseases, the level of trust in specific vaccines, and the confidence in the healthcare system (e.g. [5–7]). Modeling studies have incorporated a subset of these factors [8], such as vaccination coverage [9] and perceived disease risk [10].

More broadly, theoretical studies have modeled how the spread of disease can be affected by aspects of human behavior, particularly vaccination and social distancing behaviors [8,11–16]. Other models have examined a phenomenon known as “coupled contagion,” in which individuals can transmit not only a disease itself but also cultural factors such as vaccine adoption, disease-related fears, and (mis)information, which can in turn modulate their disease susceptibility in the simulation [10,17–19]. In real populations, health policies and other external factors can play a role in shaping vaccination cultures; two such factors are vaccine mandates, which drive vaccination uptake (even among vaccine-hesitant people), and vaccine inaccessibility, which hinders vaccine uptake (even among vaccine-confident people). Vaccine mandates have been met with opposition since their implementation in the 1800’s [20,21]. This opposition, intertwined with religious and political ideas, led to the allowance of vaccination exemptions based on medical and non-medical (e.g. religious or philosophical) reasons [22]. Though the implementation of mandatory vaccinations generally results in a drastic reduction in disease incidence and mortality [23,24], the high vaccination coverage that follows can facilitate the public perception of reduced disease severity and thus reduced vaccine necessity; this phenomenon has been observed in real populations [25,26] and incorporated into modeling studies [2,8,27]. In this vein, non-medical exemptions to mandatory vaccination have been increasing, particularly in wealthier countries where theoretical predictions suggest that belief systems can act as the main barrier to vaccination, as opposed to lack of vaccine access [28,29]. This rise in non-medical exemptions appears to have a non-trivial effect on public health, since these exemptions are correlated with the recent increase in VPD outbreaks in the United States [30,31]. However, the circumstances under which vaccine mandates might lead to increased vaccine hesitancy remain uncertain.

Even less understood is the potential association between vaccine (in)accessibility and vaccine attitudes. Vaccine accessibility issues are external pressures that negatively impact vaccination rates and coverage. Challenges to vaccine accessibility are particularly prevalent in low and middle-income countries as well as rural areas in developed countries [32,33]. For example, storage capabilities, distribution logistics, and affordability can limit the number of vaccine doses available in a specific area, and thus reduce the number of individuals who can receive a vaccine, leaving vulnerable communities at risk for a VPD outbreaks [32,34]. External factors such as limited vaccine access and vaccine mandate policies may also interact with internal factors like psychological characteristics and cultural predispositions, such as distrust in the healthcare system, potentially exacerbating the effects of low vaccine accessibility. Further, vaccination cultures can be shaped by experience with vaccines and with the disease: for example, living in a rural area could limit exposure to the disease and alter the perception of disease risk, and a lack of vaccine access for an extended period could entrench certain attitudes about vaccines in a culture. Thus, to explain the differences in vaccination outcomes and resulting disease risk across human populations, it is crucial to better understand how cultural beliefs and behaviors interact with external pressures that increase or reduce vaccination coverage.

Cultural niche construction theory describes a process in which humans modify their cultural environments—for example, their beliefs, behaviors, preferences, and social contacts—in ways that subsequently alter evolutionary pressures on the population and its culture [35]. Mathematical models of cultural niche construction have been used to explain the evolution of behaviors related to religion, fertility, and large-scale human conflict [35–40]. Since health cultures can be shaped by or influence the larger cultural landscape, the cultural niche construction framing can give insight into the cultural dynamics shaping disease risk. By using this type of model to simulate the interactions between beliefs and behaviors, we seek to understand how vaccination cultures affect vaccination coverage, as well as how vaccine-related beliefs and behaviors are affected by external forces, such as the availability of vaccines and the degree to which they are mandatory

We adapted a cultural niche construction framework to model vaccination beliefs and behaviors, incorporating the transmission of vaccination culture both from parents and from the community [9], and we used current estimates of vaccination rates and vaccine attitude frequencies obtained from various sources in the literature, including reports of Measles-Mumps-Rubella uptake from the United States [41,42], as a starting point our model. Using this model, we previously demonstrated that the overarching cultural landscape, including the likelihood of adopting vaccine hesitancy and the probability of transmitting it to one’s children, determines the equilibrium levels of vaccination coverage and vaccine hesitancy in a population. In addition, we demonstrated that the transmission of vaccine confidence and positive vaccine perception are imperative to maintaining high levels of vaccination coverage, especially when individuals preferentially choose a partner with shared vaccine beliefs. In this manuscript, we expand the scope of this model to explore how the vaccination coverage and vaccine hesitancy in a population could be affected by external forces. In particular, we focus on vaccine mandates and vaccine inaccessibility, which both lead to a decoupling of parental vaccine beliefs and their vaccination behaviors, such that vaccine mandates can increase the chances that vaccine-hesitant parents will vaccinate their children, and vaccine inaccessibility can decrease the chances that vaccine-confident parents will vaccinate their children. We quantify the population-level differences predicted by our model for populations with vaccine mandates or vaccine inaccessibility compared to a baseline population with accessible and non-mandated vaccines. Overall, we explore the effects of these external forces on the dynamics of both vaccine beliefs and vaccination coverage, providing insight into the differences between cultural development in the opposing contexts of mandates and inaccessibility.

## Methods

We build on a more general cultural niche construction framework of [9,39] to assess the effects of two external factors, vaccine mandates and vaccine accessibility, on the resulting landscape of vaccination coverage and vaccine confidence. For a population of individuals, we track the status of vaccination coverage and vaccine confidence over time; within this population, individuals mate, decide whether to vaccinate their offspring, and transmit a vaccine attitude trait. Their decision to vaccinate is influenced by their own beliefs and their vaccination states, and population trait frequencies are further modulated by vaccination-frequency-dependent cultural selection pressures. To model the effects of the external factors, we assume that vaccine mandates and inaccessibility both act to reduce the influence that internal factors, such as individual beliefs, have on vaccination behaviors.

### General Framework of the Model

Each individual in our model (depicted in **Figure 1**) has a vaccination (**V**) trait, either V**^+^** (vaccinated) or V**^−^** (unvaccinated), and an attitude (**A**) trait, either A^+^ (vaccine confident) or A^−^ (vaccine hesitant), resulting in four possible phenotypes (V^+^A^+^, V^+^A^−^, V^−^A^+^, and V^−^A^−^) that we initialize with frequencies structured to represent those of the United States: V^+^A^+^ (i.e. frequency of vaccinated, vaccine confident individuals) = 0.81, V^+^A^−^ = 0.1, V^−^A^+^ = 0.07, V^−^A^−^ = 0.02. These frequencies were estimated using reports of Measles-Mumps-Rubella vaccination rates and estimates of vaccine attitude frequencies obtained from various sources in the literature [41,42]. In each iteration, individuals mate randomly within the population. Each parental pair vaccinates their offspring with probability *B_m,n_* (i.e., vertical transmission of vaccination, with the subscript *m* denoting the vaccination trait pair and *n* denoting the attitude trait pair of the parents; see **Table 1** and **Table S1**); in general, this probability increases with each vaccinated and vaccine-confident parent. This vaccination probability is influenced by two factors: whether each of the parents are themselves vaccinated (*b_m_*), and whether each of the parents are vaccine confident or hesitant (*c_n_*). The probability that a couple vaccinates their offspring is calculated as 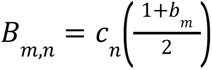, to account for the influence of both vaccination states and vaccine attitudes. We model varying levels of vaccine mandates and inaccessibility by modulating the influence that parental vaccine attitudes have on the likelihood that they vaccinate their offspring (by increasing or decreasing *c_n_*): for example, a vaccine mandate will make a vaccine-hesitant parent more likely to vaccinate their child, and vaccine inaccessibility will make a vaccine-confident parent less likely to vaccinate their child.

**Figure 1.**
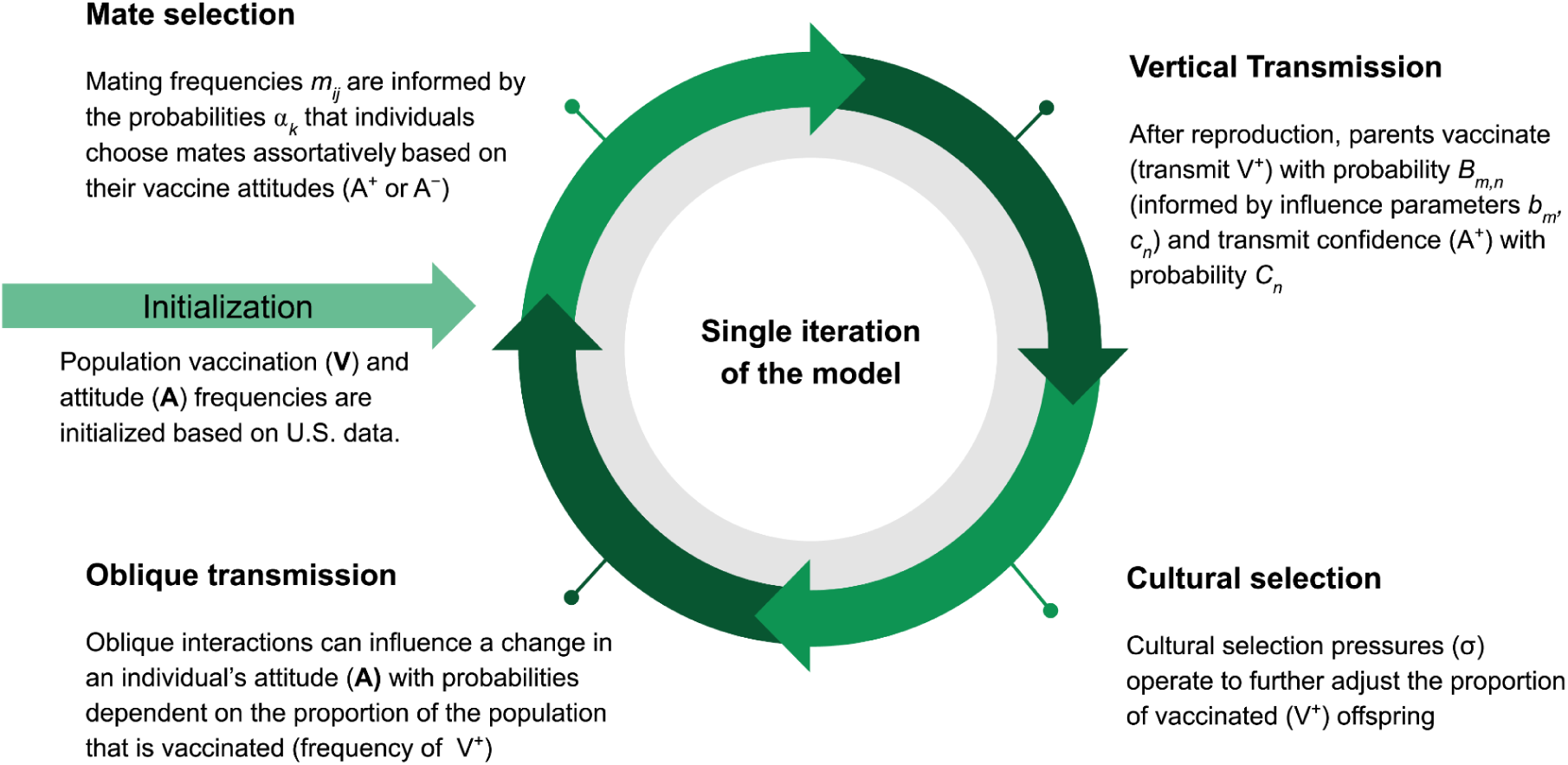
Workflow of a single iteration of the model: The schematic shows the processes within a single model iteration. The model is initialized with the phenotypic frequencies (V^+^A^+^, V^+^A^−^, V^−^A^+^, V^−^A^−^) in the population. After individuals mate and reproduce, they vertically transmit vaccination and attitude traits to their offspring. Vaccination trait frequencies are further modulated by cultural selection. Oblique transmission (cultural transmission from non-parental adults in the population) follows, which may lead offspring to alter their attitude state. (Parameters, their definitions, and baseline values are listed in **Table 1**.)

**Table 1:** List of parameters, their definitions, and baseline values.

| Parameter | Meaning | Baseline values (if applicable) |
| --- | --- | --- |
| **V** | Vaccination state ($V^+$ vaccinated, $V^-$ unvaccinated) | |
| **A** | Vaccine attitude ($A^+$ confident, $A^-$ hesitant) | |
| **m and n** | Parameter subscripts indicating traits of the mating pair (in $b_m, c_n, C_n$, and $B_{m,n}$)<br>$V^- \times V^-: m=0$; $V^- \times V^+: m=1$; $V^+ \times V^-: m=2$; $V^+ \times V^+: m=3$<br>$A^- \times A^-: n=0$; $A^- \times A^+: n=1$; $A^+ \times A^-: n=2$; $A^+ \times A^+: n=3$ | |
| **$B_{m,n}$** | Probability that parental pairs vaccinate their children, which depends upon the parents' vaccination states ($b_m$) and vaccine attitudes ($c_n$) (given in **Table S2**) | |
| **$C_n$** | Probability that parental pairs transmit vaccine confidence to their children | $C_0 = 0.01$, $C_1 = C_2 = 0.5$, $C_3 = 0.99$ |
| $b_m$ | Probability that parental pairs support offspring vaccination given their vaccination states | $b_0 = 0.01, b_1 = b_2 = 0.5, b_3 = 0.99$ |
| $c_n$ | Probability that parental pairs support offspring vaccination given their vaccine attitude | $c_0 = 0.01, c_1 = c_2 = 0.5, c_3 = 0.99$ |
| $\sigma$ | Comprehensive selection coefficient for $V^+$, dependent on the population-wide vaccination rate (see **Figure S1**) | |
| $\sigma_{\max}$ | The highest additional benefit that can be conferred by vaccination | $\sigma_{\max} = 0.1$ |

Each parental pair also transmits a vaccine attitude trait to their offspring (i.e., vertical transmission of beliefs) with vaccine confidence transmitted at probability *C_n_* and vaccine hesitancy at probability 1-*C_n_*. We set the probability of transmitting vaccine confidence to be highest for two vaccine-confident parents and lowest for two vaccine-hesitant parents (**Table 1**). For simplicity, we set the baseline confidence transmission probabilities (*C_n_*) to values reminiscent of Mendelian transmission, such that two vaccine-confident or two vaccine-hesitant parents predictably (∼100% likely) transmit their vaccine attitude, and parents with differing vaccine attitudes each have a ∼50% chance of transmitting each state: *C*_0_ near 0, *C*_1_ and *C*_2_ at 0.5, *C*_3_ near 1 **(Table 1)**. Influence parameters, *b_m_* and *c_n_*, are valued similarly and predict the probability that the couple vaccinates their children according to the equation 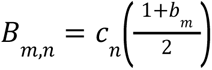, such that parents who are both vaccine confident and vaccinated are most likely to vaccinate, vaccine hesitant and unvaccinated parents are least likely to vaccinate, and parental pairs with mixed states of one or both traits will have intermediate likelihoods of vaccinating.

Each parental pair also transmits a vaccine attitude trait to their offspring (i.e., vertical transmission of beliefs) with vaccine confidence transmitted at probability *C_n_* and vaccine hesitancy at probability 1-*C_n_*. We set the probability of transmitting vaccine confidence to be highest for two vaccine-confident parents and lowest for two vaccine-hesitant parents (**Table 1**). For simplicity, we set the baseline confidence transmission probabilities (*C_n_*) to values reminiscent of Mendelian transmission, such that two vaccine-confident or two vaccine-hesitant parents predictably (∼100% likely) transmit their vaccine attitude, and parents with differing vaccine attitudes each have a ∼50% chance of transmitting each state: *C*_0_ near 0, *C*_1_ and *C*_2_ at 0.5, *C*_3_ near 1 **(Table 1)**. Influence parameters, *b_m_* and *c_n_*, are valued similarly and predict the probability that the couple vaccinates their children according to the equation 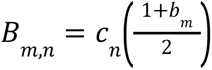, such that parents who are both vaccine confident and vaccinated are most likely to vaccinate, vaccine hesitant and unvaccinated parents are least likely to vaccinate, and parental pairs with mixed states of one or both traits will have intermediate likelihoods of vaccinating.

Next, cultural selection (σ; -1 ≤ σ ≤ 1) operates on the resulting phenotype frequencies such that the frequency of vaccination in the population is greater or less than expected given the predicted probabilities that vaccine-confident and -hesitant parents vaccinate their offspring. The proportion of vaccinated individuals in the population is multiplied by 1+σ, such that a positive σ increases the proportion of vaccinated individuals and a negative σ decreases it. This process encompasses the various factors that might make parents more or less likely to vaccinate, including the severity of the disease and the general trust in the healthcare system. Since the perceived benefit of the vaccine might vary based on the vaccination coverage in the population, we allow σ to depend on the frequency of the V^+^ trait: when the frequency of vaccination is low, the effects of the disease are more evident and individuals are more likely to vaccinate (high σ), but when the frequency of vaccination is high, the risks of the disease are masked and individuals are less likely to vaccinate (lower σ) (see **Supplement Text S1** for a more detailed explanation of how σ is calculated as a function of vaccination coverage (V^+^)). The equation relating the frequency of V^+^ and σ is given in **Figure S1**. In genetics, the selection coefficient is traditionally small (in the range of -0.1 to 0.1 [43]); at baseline in our model, we kept the maximum cultural selection coefficient at 0.1 which allowed for both positive and negative selection depending on the frequency of vaccinated individuals in the previous iteration.

Finally, oblique interactions (cultural influences from non-parental individuals) then act to further modify trait frequencies in the population. Individuals in the simulation can change their vaccine attitudes based on interactions with others and their perceptions of their surroundings. If the vaccination coverage in the population is low, we consider the negative effects of the disease to be more apparent and thus people will be less likely to adopt a vaccine-hesitant attitude, and if the vaccination coverage is high, the negative effects of the disease are prevented (amplifying the perception of the vaccine’s risks and costs, however small) and people might be more likely to become vaccine hesitant (**Figure S2**). Each subsequent iteration of the model begins with the phenotype frequencies produced at the end of the current iteration. The simulation is run until phenotype frequencies reach equilibrium (**Figure 1**, **Table 1**). For more detail see **Supplemental Text S1** and [9]. The code for the simulations is written in MATLAB and is provided at www.github.com/CreanzaLab/Vaccine-Hesitancy and http://doi.org/10.6084/m9.figshare.22493317.

### Parameterization for Mandatory Vaccination and Vaccine Inaccessibility Simulations

We hypothesize that parental vaccine attitudes influence their use of exemptions and thus levels of non-vaccination will differ based on parental attitudes under a mandated vaccination system. Therefore, we simulate the effects of mandatory vaccination by modulating the influence of a couple’s vaccine attitudes on their likelihood of vaccinating their offspring (*c*_n_); in other words, a vaccine mandate alters the influence of a couple’s vaccine attitude on their decision to vaccinate. We assume the implementation of mandates would increase vaccination in couples with at least one vaccine-hesitant individual. If vaccination exemptions are permitted, we expect that A^−^ × A^−^ couples (those with two vaccine-hesitant individuals) would be most likely to obtain exemptions, followed by mixed attitude (A^−^ × A^+^ or A^+^ × A^−^) couples, with vaccine confident couples (A^+^ × A^+^) being least likely. Hence, to model the effects of implementing a vaccine mandate, we increase attitude influence parameters from baseline values (**Table 1**) to represent two levels of mandate strictness, a strict mandate in which *c*_0_ = 0.5, *c*_1_ = *c*_2_ = 0.9, *c*_3_ = 0.99 and a less strict mandate in which *c*_0_ = 0.3, *c*_1_ = *c*_2_ = 0.7, *c*_3_ = 0.99.

Similarly, to represent a vaccine inaccessibility scenario, we reduced the influence of parental vaccine attitudes on vaccination behaviors for couples with at least one confident individual (i.e. reducing *c*_1_, *c*_2_, *c*_3_ from baseline values). In this simple representation of a vaccine-scarce environment, we assume that parents’ confidence in vaccines would have reduced influence on their ability to vaccinate their offspring, that is, their vaccine confidence does not ensure their ability to overcome vaccine inaccessibility. Hesitant couples are least likely to vaccinate their offspring regardless of vaccine availability, but couples who would likely vaccinate their offspring given the chance would have difficulty doing so due to the lack of access. We modeled two levels of vaccine inaccessibility– a somewhat inaccessible vaccine in which *c*_0_ = 0.01, *c*_1_ = *c*_2_ = 0.3, and *c*_3_ = 0.7 and an inaccessible vaccine in which *c*_0_ = 0.01, *c*_1_ = *c*_2_ = 0.1, *c*_3_ = 0.5. Assuming mixed attitude (A^−^ × A^+^ or A^+^ × A^−^) couples exhibit the most variability in their likelihood of transmitting vaccine confidence, we then examined the effect of the interaction between the maximum cultural selection coefficient (σ_max_) and mixed-attitude confidence transmission probability (*C*_1_=*C*_2_) for a scenario with baseline parameters (no active mandate and an accessible vaccine), with a less strict mandate, and with a somewhat inaccessible vaccine.

We next examined the effects of varying the transmission probability of vaccine confidence parameters for all couple types (*C*_0_, *C*_1_, *C*_2_ and *C*_3_), instead of focusing on the vaccine confidence transmission of mixed-attitude couples. We varied all *C_n_* parameters simultaneously within a specified range of values (**Table S3**) across different levels of mandate strictness and vaccine inaccessibility. As before, we varied these parameters in conjunction with the maximum cultural selection coefficient σ_max_.

## Results

### Mandatory Vaccination and Vaccine Inaccessibility

We examined the effect of the interaction between the maximum cultural selection coefficient (σ_max_) and confidence transmission probability of mixed-attitude couples (A^−^ × A^+^ and A^+^ × A^−^; *C*_1_=*C*_2_) (**Figure 2**). Modeling the effects of a vaccine mandate reveals a decoupling of vaccination coverage and vaccine confidence trajectories when parents are more likely to transmit vaccine hesitancy (**Figure 2C-D**). Even when vaccine confidence is very low (specifically at mixed-trait couple confidence transmission probabilities below 0.5; red region in **Figure 2D**), vaccination coverage is higher with a less strict mandate implemented than without a mandate (compare **Figure 2C-D** to **Figure 2A-B; Supplemental Table S4**). However, the leniency of the mandate in **Figure 2C-D** means that many vaccine-hesitant couples can obtain an exemption, and vaccination coverage remains lower when vaccine hesitancy is common. This suggests that an external pressure to vaccinate helps overcome the opposing cultural pressure imposed by hesitancy in the population, but a mandate would have to be stricter to achieve herd immunity in a predominantly vaccine-hesitant population.

**Figure 2:**
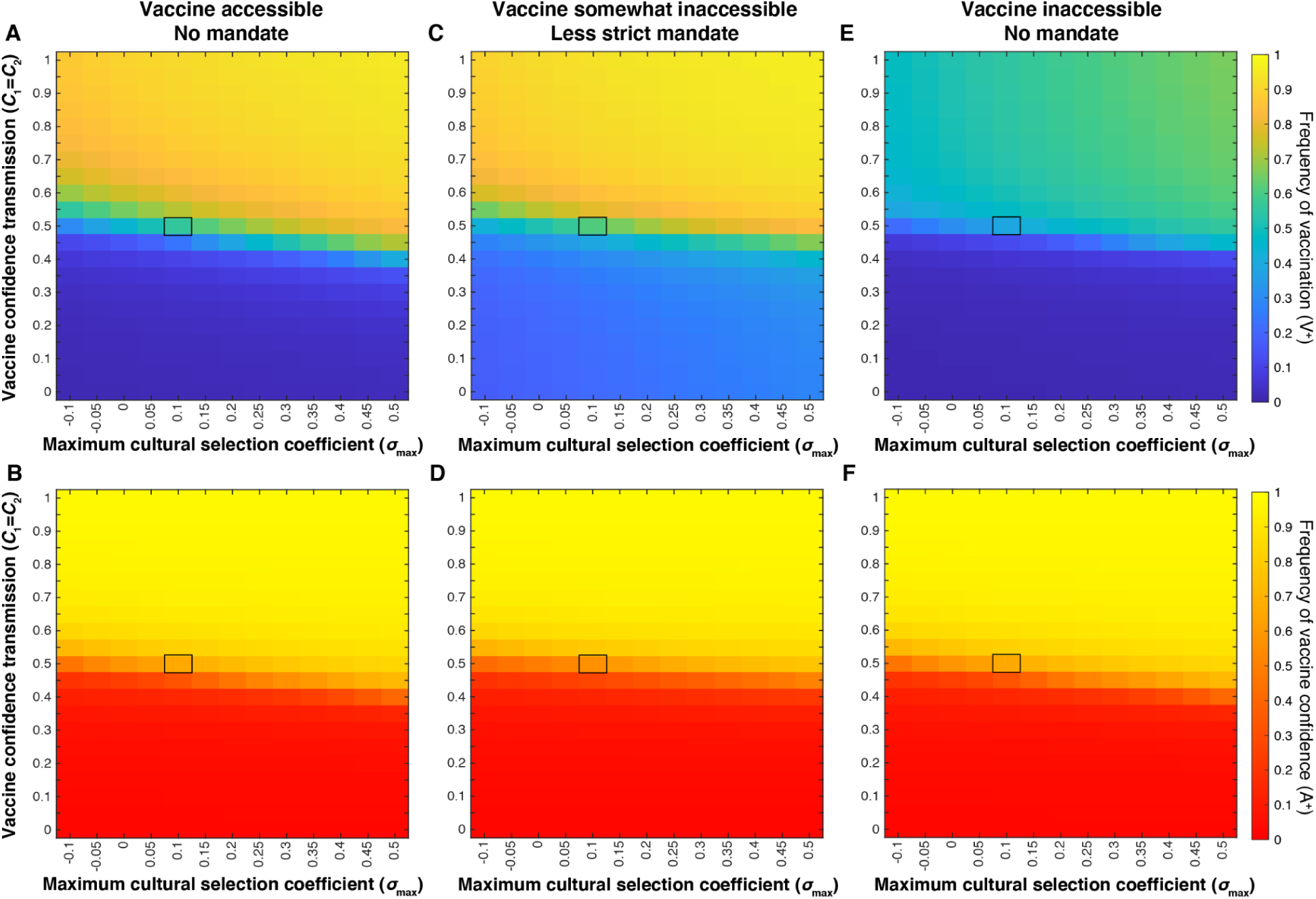
External factors (vaccine mandates and vaccine scarcity) decouple equilibrium levels of vaccine confidence from vaccination coverage. Heatmaps showing equilibrium vaccine coverage and vaccine confidence levels with an accessible vaccine and no active mandate (**A, B**), with an accessible vaccine and a less strict mandate (**C, D**) and an environment with vaccines somewhat inaccessible (**E,F**). Assuming mixed-attitude couples might have the most variability in their likelihood of transmitting vaccine confidence to their offspring, we vary *C*_1_ = *C*_2_ (confidence transmission probability of mixed-attitude couples) on the vertical axis, and maximum selection coefficient σ_max_ (indicative of the perceived value of vaccinating offspring) on the horizontal axis. A less strict mandate (**C, D**) is modeled by *c*_0_ = 0.3, *c*_1_ = *c*_2_ = 0.7, *c*_3_ = 0.99; vaccine inaccessibility (**E, F**) is modeled by *c*_0_ = 0.01, *c*_1_ = *c*_2_ = 0.3, *c*_3_ = 0.7. Unspecified parameters are given in **Table 1**. These simulations show an inverse correlation between vaccination coverage and vaccine confidence at *C_n_* < 0.5 under a less strict mandate, and *C_n_* > 0.5 when vaccine access is limited. Baseline conditions (**Table 1**) are highlighted by black boxes in each heatmap. To facilitate comparisons between panels, the mean and median for the section of the heatmaps with *C*_1_ = *C*_2_ < 0.5 are presented in **Supplemental Table S4**.

When vaccines were somewhat inaccessible, vaccination coverage was noticeably reduced overall, while vaccine confidence increased slightly across the parameter space. Juxtaposed with the mandate scenario **(Figure 2C-D)**, our vaccine scarcity models produce an opposite deviation of vaccination coverage from vaccine confidence levels: when vaccines are mandated, we observe increased vaccination coverage in low-confidence environments, and when vaccines are inaccessible, we observe lower than expected vaccination coverage (<50%) in a predominantly vaccine-confident environment (>90%) (**Figure 2**).

### Mandatory vaccination may increase vaccination coverage at the expense of confidence, while vaccine inaccessibility promotes confidence

In the three scenarios examined thus far—baseline (no mandate and accessible vaccines), a less strict mandate, and somewhat inaccessible vaccines—most of the variability in equilibrium frequencies across the parameter space occurs at confidence transmission levels between *C*_1_ = *C*_2_ = 0.4 to 0.6 (**Figure 2**). This threshold region separates definitively higher and definitively lower vaccination coverage and vaccine confidence outcomes. The effect of actual and perceived vaccine fitness (σ) is also most noticeable in this region of the heatmap: as cultural selection for vaccination increases at any fixed probability of confidence transmission, vaccination coverage and vaccine confidence levels at equilibrium are increased. Changes in vaccination and confidence frequencies are not independent of each other, as these effects are the consequence of changes in phenotypic frequencies. Therefore, for each scenario, we plotted the temporal dynamics of each phenotype (**VA**) and the vaccination (V^+^) and confidence (A^+^) traits at baseline parameter values (**Figure 3**), then calculated the difference in frequency from baseline equilibrium (**Table 2**). With an accessible vaccine that is not mandated (**Figure 3A**, **Table 2**), the phenotype frequencies of the system equilibrate generally with either vaccinated and vaccine confident (V^+^A^+^) or unvaccinated and vaccine hesitant (V^−^A^−^) individuals most abundant (**Figure 3A**, **Table 2**). Though these two phenotypes remain the most abundant when a less strict vaccine mandate is implemented, the equilibrium frequency of vaccinated but vaccine-hesitant individuals (V^+^A^−^) is greatly increased compared to baseline (**Figure 3B**, **Table 2**). Interestingly, a mandate also results in a higher frequency of unvaccinated and vaccine-hesitant individuals (V^−^A^−^), while reducing vaccinated and vaccine-confident individuals (V^+^A^+^) in the population. Vaccine inaccessibility, on the other hand, resulted in approximately double the frequency of unvaccinated but vaccine-hesitant (V^−^A^+^) individuals. In summary, compared to baseline outcomes, implementation of a mandate increases vaccination coverage at the expense of confidence by driving vaccination in hesitant individuals, and vaccine inaccessibility promotes confidence despite low vaccination coverage by driving confidence in unvaccinated individuals.

**Figure 3:**
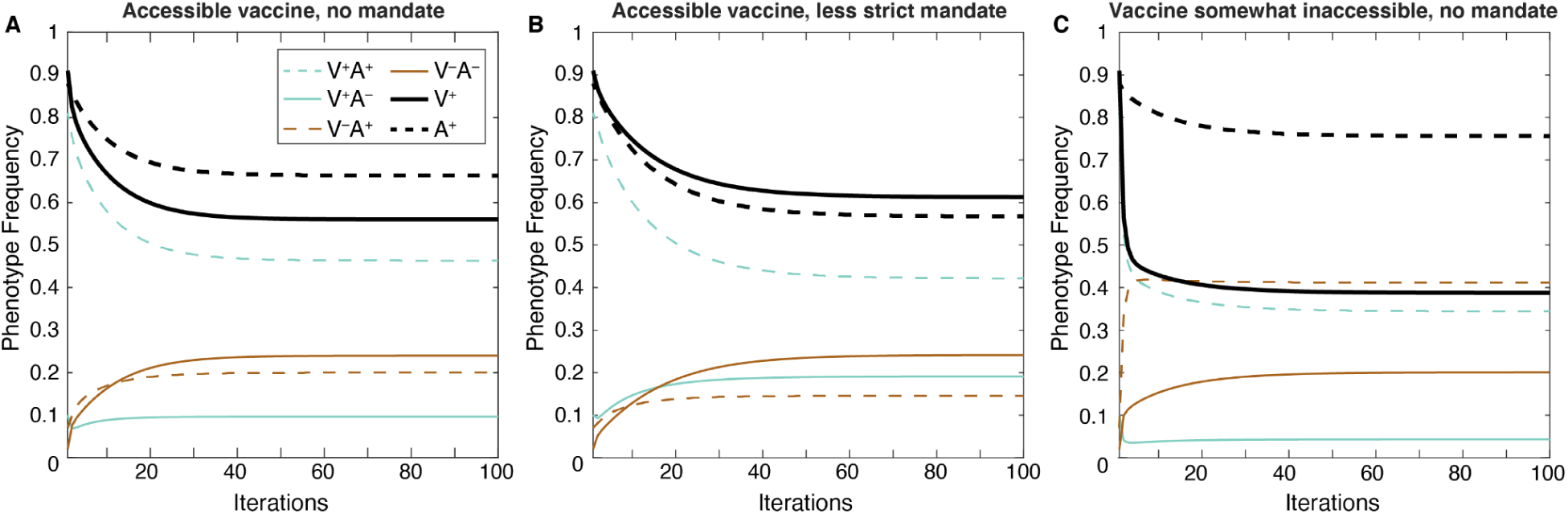
Vaccine mandates and inaccessibility drive different distributions of both vaccination coverage and vaccine confidence. Phenotype and trait frequencies are plotted over 100 model iterations. Compared to baseline transmission levels (panel **A**, parameter values given in **Table 1**), a less strict vaccine mandate (*c*_0_ = 0.3, *c*_1_ = *c*_2_ = 0.7, *c*_3_ = 0.99; panel **B**) leads to increased vaccination coverage at equilibrium (black line) but decreased vaccine confidence levels (magenta line). In contrast, when a vaccine is somewhat difficult to access (*c*_0_ = 0.01, *c*_1_ = *c*_2_ = 0.3, and *c*_3_ = 0.7; panel **C**), vaccination coverage is lower than in panel **A** but vaccine confidence is higher. The specific simulations shown here are highlighted with black rectangles on the heatmaps in Figure 2.

**Table 2:** Change from Baseline Equilibrium Frequencies. Final equilibrium frequencies for baseline, a less strict vaccine mandate, and a somewhat inaccessible vaccine are shown along with the percent difference from baseline frequencies. Colored lines in the first row correspond to the line colors in **Figure 3**. Negative changes are indicated by a red downward pointing triangle; positive changes are indicated by green upward pointing triangle. A vaccine mandate leads to increased vaccination among vaccine-hesitant individuals, and vaccine inaccessibility leads to decreased vaccination and increased vaccine confidence among unvaccinated individuals.

| Phenotype |  | V+A+ — — | V+A- — | V-A+ — — | V-A- — | V+ — | A+ — — |
| --- | --- | --- | --- | --- | --- | --- | --- |
| Baseline Equilibrium Frequencies |  | 46.34% | 9.66% | 20% | 23.99% | 56.01% | 66.34% |
| Percent Diff. from Baseline | Mandate | 42.17%<br>(-9%) ▼ | 19.11%<br>(98%) ▲ | 14.58%<br>(-27%) ▼ | 24.23%<br>(1%) ▲ | 61.29%<br>(9%) ▲ | 56.75%<br>(-14%) ▼ |
|  | Inaccessibility | 34.43%<br>(-26%) ▼ | 4.33%<br>(-55%) ▼ | 41.16%<br>(106%) ▲ | 20.07%<br>(-16%) ▼ | 38.77%<br>(-31%) ▼ | 75.60%<br>(14%) ▲ |

### Vaccination and confidence frequencies are more variable when offspring beliefs are more likely to differ from their parents’ beliefs

The clear disjunction between higher and lower vaccination (V^+^) and vaccine confidence (A^+^) frequencies observed in **Figure 2** is not observed when the probability of confidence transmission is modulated for all couples (**Figure 4**). When mixed-attitude couples transmit confidence to their offspring at high (*C*_1_ = *C*_2_ > 0.5) or low (*C*_1_ = *C*_2_ < 0.4) probabilities, which skews population attitude frequencies to either highly confident or highly hesitant, the subsequent offspring are more likely to vaccinate (in a confident population) or not vaccinate (in a hesitant population) (**Figure 2**). Similarly, if all couple types are transmitting confidence at lower probabilities or higher probabilities (i.e. *C*_0_, *C*_1_, *C*_2_, and *C*_3_ are all lower or higher, respectively), vaccination frequencies will equilibrate at either lower levels or higher levels (**Figure 4A**). However, if all couples are transmitting confidence at mid-range probabilities (or *C*_1_ and *C*_2_ are closer to 0.5), the population equilibrates at more polymorphic frequencies, that is, both forms of each trait coexist in the population at moderate frequencies.

**Figure 4:**
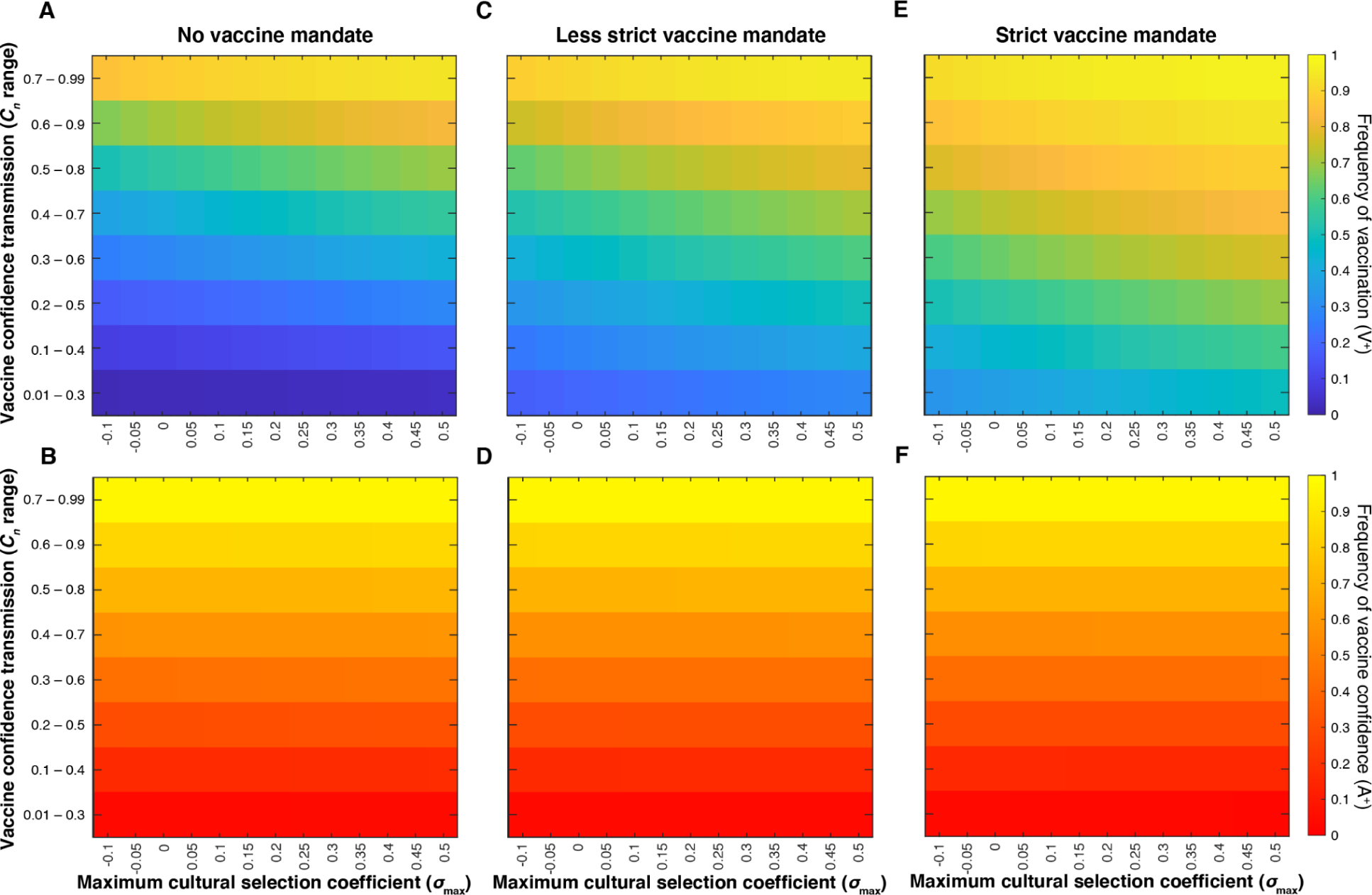
Increasing mandate strictness and increased cultural selection drive vaccination coverage. Heatmaps showing final vaccination coverage (**A, C, E**) and corresponding vaccine confidence (**B, D, F**) after 100 time-steps while simultaneously varying all confidence transmission probabilities (*C_n_*; vertical axis) and maximum selection coefficient (σ_max_; horizontal axis). We show an accessible vaccine with no mandate (*c*_0_= 0.01, *c*_1_= *c*_2_ = 0.5, *c*_3_ = 0.99) (**A, B**), a less strict mandate (*c*_0_ = 0.3, *c*_1_ = *c*_2_ = 0.7, *c*_3_ = 0.99) (**C, D**), and a strict mandate (*c*_0_ = 0.5, *c*_1_ = *c*_2_ = 0.9, *c*_3_ = 0.99) (**E,F**). *C_n_* values are set within the range indicated on the vertical axis with *C*_0_ taking the lowest value, *C*_1_ and *C*_2_ taking intermediate values, and *C_3_* taking the highest value (**Table S3**).

Equilibrium vaccination coverage increases as cultural selection for vaccination increases in both mandated vaccines (**Figure 4C, E**) and vaccine inaccessibility scenarios (**Figure 5C**, E); confidence frequencies remain more consistent across the range of cultural selection pressures (**Figure 4D**, F, **Figure 5D**, F). When we model an increase in vaccine mandate strictness (increased difficulty in obtaining exemptions), vaccination frequencies are increased (**Figure 4C**, E). On the other hand, greater degrees of inaccessibility lead to larger reductions in vaccination coverage (**Figure 5C**, E), and lower coverage occurs despite higher levels of vaccine confidence

**Figure 5:**
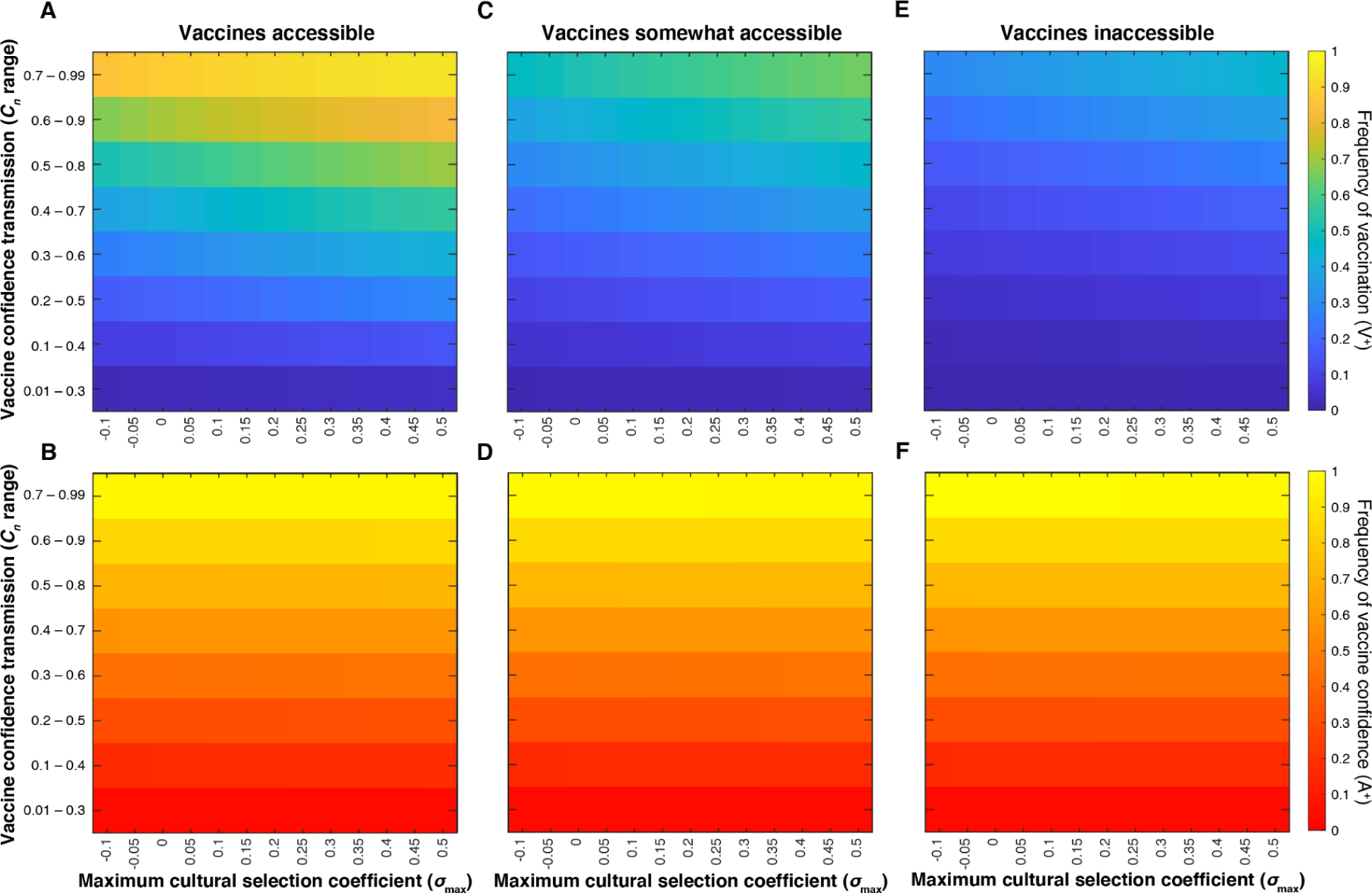
Vaccine inaccessibility reduces vaccination coverage despite high levels of vaccine confidence. Heatmaps showing final vaccination coverage (**A, C, E**) and corresponding vaccine confidence (**B, D, F**) after 100 time-steps while simultaneously varying all confidence transmission probabilities (*C_n_*; vertical axis) and maximum selection coefficient (σ_max_; horizontal axis). *C_n_* values are set within the range indicated on the vertical axis with *C_0_* taking the lowest value, *C*_1_ and *C*_2_ taking intermediate values, and *C_3_* taking the highest value (**Table S3**). We simulate an accessible vaccine and no mandate (*c*_0_= 0.01, *c*_1_= *c*_2_ = 0.5, *c*_3_ = 0.99) (**A, B**), a somewhat inaccessible vaccine (*c*_0_ = 0.01, *c*_1_ = *c*_2_ = 0.3, and *c*_3_ = 0.7) (**C, D**) and an inaccessible vaccine (*c*_0_ = 0.01, *c*_1_ = *c*_2_ = 0.1, *c*_3_ = 0.5) (**E,F**).

### Changing the relationship between vaccination coverage and cultural selection can alter vaccination behavior when the vaccine is accessible

In the previous analyses, we assumed that the cultural selection for vaccination would begin to decrease from its maximum value as members of a population with widespread vaccination coverage (exceeding 70% vaccination coverage, see **Figure S1**) might perceive a reduced cost of the disease and thus a reduced pressure to vaccinate their children. To assess the robustness of our model to different relationships between vaccination coverage and cultural selection pressures, for example representing variations in herd immunity criteria or in parent priorities, we tested the same simulations with multiple cultural selection functions. We examined the interaction between mixed-attitude pair confidence transmission probability (*C*_1_ = *C*_2_) and a range of maximum cultural selection coefficients (σ_max_) for these different cultural selection functions (shown in **Figure S3A** for σ_max_ = 0.1). In line with cultural selection acting primarily on the vaccination trait, most of the differences among the cultural selection functions are observed in the vaccination equilibrium frequencies and not the confidence equilibrium frequencies, particularly when no mandates or less strict mandates are imposed (**Figure S3B-C**). Compared to the baseline function used in **Figures 2-5** (also shown in **Figure S3, column 3**), when we define “herd immunity” as being achieved at a reduced vaccination coverage level (corresponding to when σ begins to decrease), vaccination coverage is reduced most noticeably at the intersection of low values of σ_max_ and high values of mixed-attitude pair confidence transmission (**Figure S3B-C, column 4**). When the level required for herd immunity is increased, vaccination coverage is increased in this low σ_max_ high confidence transmission area of the heatmap (**Figure S3B-C, column 2**). The overall patterns we observed with the original cultural selection function are robust to the particular function we used. At higher values of *C*_1_=*C*_2_ and σ_max_ (top right corner of the heat maps), vaccination coverage was reduced when the σ function was more negatively correlated with vaccination coverage (columns 3 through 6 in **Figure S3**). In addition, we observed that the largest differences in vaccine coverage between cultural selection (σ) functions occurred when vaccines were accessible and there were no mandates (**Figure S3B**); differences in the cultural selection function had less of an effect on vaccination coverage when a less strict mandate was imposed (**Figure S3C**), and had little effect on vaccination coverage when vaccines were inaccessible (**Figure S3D**). The least variation is observed when vaccines are inaccessible: across confidence frequencies, the cultural selection function did not meaningfully alter the equilibrium vaccination coverage or vaccine confidence (**Figure S3D**). This result is intuitive, since most differences between cultural selection functions occur in regions of high vaccination coverage, and the simulations with inaccessible vaccines do not not lead to high vaccination coverage for any parameter combination.

## Discussion

Here, we build on the cultural niche construction framework proposed by [9] to model the cultural spread of vaccine attitudes and vaccination behavior in the presence of external forces imposed by two scenarios: vaccine mandates and vaccine inaccessibility. Multiple factors influence an individual’s vaccine-related beliefs and a couple’s decision to vaccinate their offspring, including their own vaccination status and their perception of the relative risks of the disease and the vaccine. As such, it is important that we understand how public health policies, such as vaccine mandates and barriers to vaccination, such as geography or affordability, can shape vaccination cultures and thus affect public health. Using a cultural niche construction approach allows us to explore the effects of the interplay between external forces and cultural factors providing further insight into how vaccination cultures are formed, maintained, and evolve.

With our initial model [9], we showed that when population traits are at or near an equilibrium, we can infer that a population with high vaccination coverage will have low rates of vaccine hesitancy and vice versa. However, when there are external pressures as modeled here, such as increased pressure to vaccinate or difficulty in acquiring vaccination exemptions, an undercurrent of vaccine hesitancy can persist in a relatively well-vaccinated population, potentially limiting the adoption of newly introduced vaccines. This possibly contributes to the unexpected lag in uptake of newer vaccines, such as the COVID or HPV vaccines, in communities with otherwise high vaccination rates [44–46]. The perceived increase in hesitancy surrounding new vaccines may actually be existing vaccine hesitancy becoming apparent. In addition, “fence sitters”, those who have not made a firm stance regarding vaccines and thus could be more influenced by targeted campaigns [42], may develop higher levels of uncertainty about new vaccines than their parents had about existing ones.

In contrast to the effect of vaccine mandates, by modeling vaccine inaccessibility we illustrate another important pattern: reduced vaccination coverage in a vaccine-confident culture. In a vaccine-scarce environment, an individual’s attitude regarding vaccines has less influence on vaccination behavior due to the barrier imposed by resource availability. As a result, a population may be undervaccinated despite holding vaccine-affirming beliefs. In addition, a health culture previously shaped by vaccine inaccessibility could potentially ingrain specific behavioral practices (for example, visiting the doctor only when a child is sick and not for a regular vaccine schedule) that are not easily modified even if vaccines become more readily available. These vaccine scarcity scenarios are most likely to exist in low- and middle-income countries in which vaccine acquisition, storage and/or distribution resources are insufficient [47–49] whereas the opposite scenario (low vaccine confidence–high vaccination coverage) after vaccine mandates is most common in developed nations [50]. In summary, we find that vaccine mandates can result in high vaccination coverage even in a culture of hesitancy, and that lack of access to vaccines can produce the inverse: low vaccination coverage in a culture of confidence.

It is difficult, as with any system, to fully capture the complex reality of vaccine hesitancy and vaccination behavior with a mathematical model. Caveats of this model include the lack of empirical data to inform how we model the influence of vaccine confidence on vaccination behaviors in the face of mandates or vaccine inaccessibility. A potential limitation of modeling cultural selection as a function of vaccination frequency is that the potential values of the cultural selection coefficient could be restricted when vaccines are scarce. The shape of our default cultural selection coefficient function (**Figure S1**) assumes less variability in the perceived benefit of vaccination at low levels of vaccination coverage. Since the major limiting factor of vaccination frequency in a vaccine-scarce environment could be vaccine availability, rather than vaccination behavior, the higher limit of population vaccination frequency is externally reduced based on vaccine supply, thus biasing vaccine perception and vaccination selection in our model to be close to σ_max_ (highest selection for vaccination). It is possible that the trajectory of cultural selection for vaccination may change when vaccines are scarce: perhaps patterns of perceptions change with the knowledge of vaccine (un)availability [51], thus potentially providing an avenue for further exploration with this model. Though not explicitly discussed, we test scenarios in which the selection coefficient is more sensitive to changes in lower vaccination frequencies, which might be relevant when vaccines are inaccessible (**Figure S3 Column 5-6**). Compared to our baseline selection function (**Figure S3 Column 3**), the change in sensitivity does give rise to slightly lower levels of vaccination coverage at equilibrium when confidence transmission by mixed-trait couples is higher.

In addition, our model simplifies the process of human population turnover with discrete generations; in reality, of course, population turnover is asynchronous and multiple generations can have cultural interactions with one another [9]. However, this simple model is able to demonstrate interesting scenarios that confirm the importance of understanding the culture of the communities in which public health policies act, and how the cultural landscape might affect specific outcomes. A community is most protected from VPD outbreaks if two conditions are met: vaccination coverage achieves or exceeds herd immunity levels, and future vaccinations are not threatened by underlying vaccine hesitancy. The effects that we observe as a result of varying the cultural selection function suggest that an “unwavering” (positive) perception of vaccination is better for maintaining higher levels of vaccination coverage than one that varies with vaccination coverage. This highlights a significant issue in increasing vaccination in the absence of (severe) disease as perceptions are shaped by experience of both the disease and measures used to address the disease. Since increasing vaccination coverage might require different strategies than increasing confidence, we encourage public health policymakers to consider both beliefs and behavior patterns in their outreach efforts and information campaigns.

The results of our simulations are congruent to those observed in other behavior change model studies [16]. For example, Epstein *et al*. [17] demonstrated using a “triple contagion” model, in which a disease, fear of a disease, and fear of a vaccine can each be transmitted between individuals, that high vaccination coverage may be achieved when fear of a vaccine is low and fear of the disease is high. Though our model uses different methods of transmission, we arrive at similar conclusions; for example, our model predicts higher vaccination coverage when the cultural selection coefficient is high, suggesting a higher perceived value of vaccination (and thus lower fear of the vaccine). Similarly, faster spread of vaccine fear in the Epstein *et al.* study could be interpreted similarly to higher probabilities of transmitting vaccine hesitancy (lower *C*_1_ = *C*_2_ values) in our model, and we also observe reduced vaccination coverage in these scenarios.

Another external factor that can affect these dynamics, but has not been explored in this study, is the occurrence of a pandemic and the introduction of a novel vaccine. The COVID-19 pandemic appears to have had complex ramifications on attitudes toward other vaccines. For example, a global survey assessing caregiver willingness to vaccinate their children against influenza showed that changes in caregiver risk perception due to COVID-19 and concerns that their children may have contracted COVID-19 resulted in a significant upward shift in caregiver’s plan to vaccinate against influenza following the pandemic – approximately 29% of caregivers who had not vaccinated their children in the previous season (2019-2020) planned to vaccinate in the next [52]. A recent report on kindergarten vaccination rates in Tennessee illustrated a temporal correlation between the pandemic and an increase in the use of non-medical, particularly religious, vaccine exemptions [53]. The same report also noted that the barriers to obtaining routine medical care and administrative challenges in schools during the COVID-19 pandemic contributed to increased vaccine-hesitant behavior such as modifying vaccine schedules.

Additionally, an experimental study of the effects of COVID-19 vaccine scarcity [54] found that vaccine scarcity could decrease the willingness to vaccinate, but it did not, however, affect the perception of risk or protection associated with the vaccine. Though the perceived risk in our model is modulated according to vaccination frequency (that is, in our model, perceptions are modulated by vaccination coverage), our simulations reveal an intuitively similar pattern: vaccination is reduced overall when vaccines are scarce. However, while perception may be modulated in our model, we do observe an increase in vaccine confidence under conditions that result in low vaccination coverage. This is in line with the findings of Pereira *et al.* [54] as vaccine-confident individuals may choose to forgo vaccinations for the benefit of others if resources are limited, such as when the COVID-19 vaccine was relatively inaccessible when it was initially released, while still maintaining (and transmitting) their vaccine beliefs. In contrast to our model, which focuses on established childhood vaccines, the Pereira *et al.* study focused on adult vaccination with the novel COVID-19 vaccine. The differences between the vaccine target populations (e.g. child vs. adult) and the interacting individual values (e.g. compassion for higher-risk individuals in foregoing one’s own vaccinations when vaccines are scarce) may produce differing dynamics requiring different public health approaches.

In sum, our model shows, in both mandate and inaccessibility scenarios, that the probability of transmitting vaccine-positive attitudes is a strong predictor of whether future vaccination coverage is high or low (**Figures 2**, **4-5**). We also demonstrate that vaccine efficacy and perceived value are important to maintaining sufficient levels of vaccination coverage, especially if vaccine confidence is not being robustly transmitted (or maintained in adulthood), regardless of vaccination scenario (**Figures 2**, 4-5). Thus, our model demonstrates the importance of clear and accurate communication about vaccines even when vaccination is mandatory and resulting coverage is high, to reduce the spread of inaccurate information that can foster vaccine hesitancy and hinder the uptake of future vaccines. Taken together, the results our model suggest that combatting low or declining vaccine uptake would take a sophisticated approach that targets the physical vaccination behavior (availability and mandates) while simultaneously addressing a population’s constantly evolving vaccine perceptions.

## Data Availability

All data and code are available online at https://github.com/CreanzaLab/Vaccine-Hesitancy

## Notes

### Competing Interest Statement

The authors have declared no competing interest.

### Funding Statement

This study was funded by Vanderbilt University and the John Templeton Foundation.

### Summary of Updates

We modified the manuscript in response to feedback; in particular, we clarified the text, added the results of additional simulations to assess the influence of the cultural selection function on the model's results, and edited Figure 3 to be more printer- and colorblind-friendly.

