## Supplementary Materials for "Internal and external factors affecting vaccination coverage: modeling the interactions between vaccine hesitancy, accessibility, and mandates"

**Text S1: Detailed Methods**

Here, we expand on the model proposed in [26]. We consider two cultural traits: a vaccination trait (**V**), and a vaccine attitude trait (**A**). Each individual can take one of two possible states for each trait, $V^+$ (vaccinated) or $V^-$ (unvaccinated) and $A^+$ (vaccine confident) or $A^-$ (vaccine hesitant), respectively. This results in four possible phenotypes: $V^+A^+$ (type 1: vaccinated and confident), $V^+A^-$ (type 2: vaccinated and hesitant), $V^-A^+$ (type 3: unvaccinated and confident), and $V^-A^-$ (type 4: unvaccinated and hesitant), whose frequencies in the population are denoted by $x_1$, $x_2$, $x_3$, and $x_4$, respectively, with $\sum_{i=1}^{4} x_i = 1$. (See **Table S1** for subscript assignments).

The four phenotypes described produce sixteen possible mating pairs. The mating frequency, $m_{i,j}$ indicates the frequency of a mating between a parent of type $i$ and the second parent of type $j$ where $i, j=\{1, 2, 3, 4\}$ (**Table S1**); for example, $m_{1,3}$ represents the mating frequency of $V^+A^+$ ($x_1$) and $V^-A^+$ ($x_3$). In this manuscript, we assume random mating, therefore individuals of different phenotypes mate with one another at a rate equal to the product of their frequencies.

Since the two traits (**A** and **V**) are transmitted vertically, for each phenotype we specify the probability that the mating produces an offspring of phenotype ($V^+A^+$). The vaccine confidence trait ($A^+$) is transmitted with probability $C_n$, and the vaccine hesitancy trait ($A^-$) is transmitted with probability $1-C_n$ (for $n=\{0, 1, 2, 3\}$ as shown in **Table S2**). If $C_0 = 0$, two $A^-$ parents will always produce $A^-$ offspring, and if $C_3 = 1$, two $A^+$ parents will always produce $A^+$ offspring. However, if $C_0 > 0$, two $A^-$ parents can produce $A^+$ offspring at some probability, and similarly if $C_3 < 1$, two $A^+$ parents can produce $A^-$ offspring with some probability.

Transmission of vaccination ($V^+$ with probability $B_{m,n}$ for $m, n=\{0, 1, 2, 3\}$; **Table S2**) is more complex, since parents' vaccine attitudes (**A**), in addition to their own vaccination states (**V**), can influence their behavior in vaccinating their offspring via a set of "influence parameters" that inform vaccination probabilities. The probability that each mating pair produces an offspring with the $V^+$ trait (i.e. vaccinates their offspring) is a scaled product of the influence of parental attitudes ($c_n$ for $n=\{0, 1, 2, 3\}$) and the influence of parental vaccination states ($b_m$ for $m = \{0, 1, 2, 3\}$ ) (**Table S2**). For example, for mating pair $V^+A^+ \times V^+A^-$, their combined vaccination states ($V^+$

× $V^+$) will influence vaccination behavior by $b_3$, and their combined attitude states, ($A^+ \times A^-$), will influence vaccination behavior by $c_2$. Therefore, a $V^+A^+ \times V^+A^-$ mating will produce a $V^+$ offspring with probability $B_{3,2} = c_2\left(\frac{1+b_3}{2}\right)$; this pair will also produce an $A^+$ offspring with probability $C_2$ based on their combined attitude states.

**Table S1: Presence (+) and absence (–) subscript assignments.** Demonstrating the trait presence (+) and absence (–) combinations associated with m, n subscripts. For example, the + × – combinations is associated with m and n subscript value 2: an $A^+ \times A^-$ pairing transmits $A^+$ at probability $C_2$. This rule applies to parameters $C_n$, $b_m$, $B_{m,n}$, $c_n$, as shown in **Table S2**.

| Subscript Value (*m*, *n*; e.g. $b_m$, $C_n$) | Associated Pairing (e.g. V × V, A × A) |
|---|---|
| 0 | – × – |
| 1 | – × + |
| 2 | + × – |
| 3 | + × + |

Transmission and influence probabilities are constant throughout a single simulation, with values ranging from 0 to 1. At baseline settings, the influence parameters $b_m$ and $c_n$, and the transmission parameter $C_n$ would take the values indicated in **Table 1**. In our model, vaccination probabilities are structured such that a couple's vaccine beliefs have a greater influence ($c_n$) on their likelihood of vaccinating their offspring than their own vaccination status ($b_n$). Therefore, offspring vaccination is guaranteed at some probability only if $c_n > 0$. We implement vaccine mandates and vaccine inaccessibility by modulating the influence of vaccine attitudes ($c_n$). We increase the influence parameter values of couples with at least one vaccine hesitant individual ($c_0$: $A^- \times A^-$, $c_1$: $A^- \times A^+$, $c_2$: $A^+ \times A^-$ ) to model a vaccine mandates or decreasing influence parameter values of couples with at least one vaccine confident individual ($c_3$: $A^+ \times A^+$, $c_2$: $A^+ \times A^-$, $c_1$: $A^- \times A^+$) to model vaccine inaccessibility. In other words, a vaccine mandate will make a vaccine-hesitant parent more likely to vaccinate their child, and vaccine inaccessibility will make a vaccine-confident parent less likely to vaccinate their child.

The cultural selection on vaccination is given by the parameter σ. After vertical cultural transmission has occurred, the frequency of the $V^+A^+$ and $V^+A^-$ phenotypes are multiplied by 1+σ. This parameter modulates whether there are more or fewer vaccinated individuals than expected: in other words, when $\sigma>0$, vaccinated individuals are more common in a set of offspring than would be expected strictly by parental beliefs and vaccination statuses. This cultural selection coefficient is structured to encompass both biological fitness and cultural selection pressures, including perceived risks or benefits of the vaccine itself, personal cost-benefit analyses of preventative health behaviors, and the structural or societal-level factors influencing vaccination rates [1,2]. Under the assumption that effects of herd immunity may lead to a reduction in vaccination behaviors—for example, the belief that vaccines are unnecessary when most others are vaccinated [3]—the cultural selection coefficient function in our model is vaccine-frequency-dependent. We calculate $\sigma$ in each timestep as a function of the current vaccination coverage (frequency of $V^+$, i.e. $x_1 + x_2$), and in each simulation we specify $\sigma_{max}$ as the maximum cultural selection pressure for getting vaccinated ($-1 \leq \sigma_{max} \leq 1$) (see the cultural selection coefficient function in **Figure S1**). To incorporate this relationship into the model, we constructed a function by defining our assumptions (incorporating evolutionary game theory, e.g. the "free rider" problem) and then choosing curves with a trajectories that met pre-specified conditions: with unvaccinated individuals holding baseline fitness at 0, we assume that when vaccination coverage is low, the real and perceived benefits of vaccination are highest, and thus, the cultural selection pressure is near $\sigma_{max}$, however, as vaccination coverage increases toward the level of herd immunity, the perceived benefits of vaccination decrease, represented as a reduction in the cultural selection pressure [4].

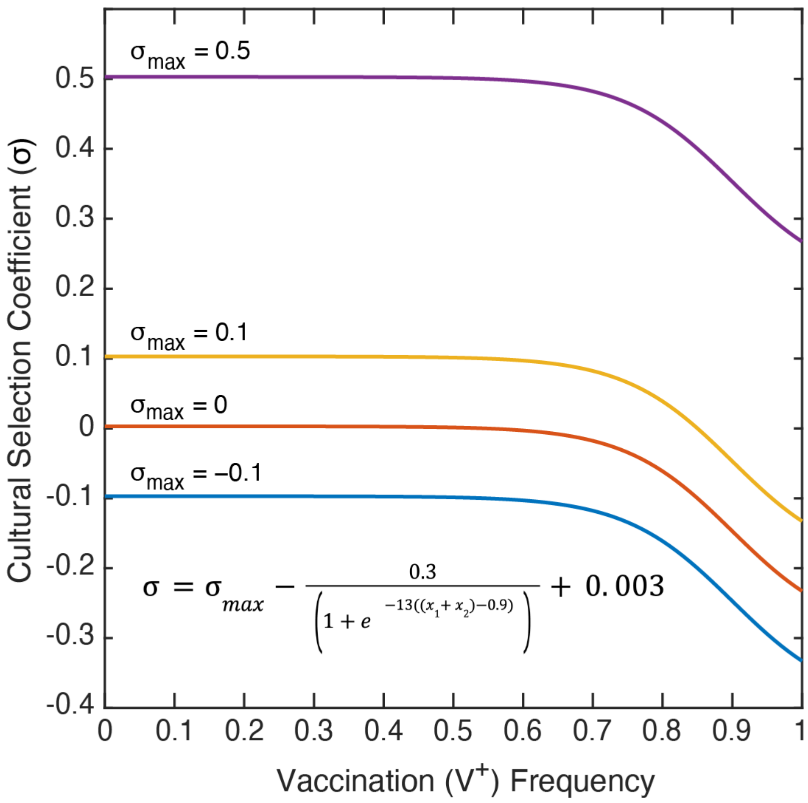


**Figure S1: Cultural selection coefficient function**. The cultural selection coefficient considers both health and non-health related effects, and the function was constructed by fitting a curve to specified conditions. The selection coefficient (σ; vertical-axis) is dependent on the frequency of vaccinated individuals ($V^+$) in the population (horizontal-axis). $\sigma_{max}$ is the maximum cultural selection coefficient associated with being vaccinated. Perceived vaccine benefit is reduced as vaccination coverage increases, since the negative effects of the disease will be less apparent.

The model incorporates a second phase with oblique cultural transmission (i.e. influence from non-parental adults), in which individuals can change their inherited vaccine attitudes (**A**) due to influence from other adults in the population. There are two probabilities associated with attitude modulation: the probability that an vaccine hesitant ($A^-$) individual adopts the vaccine confident ($A^+$) state ($A^-$ to $A^+$ transition probability, given by $A_{\rightarrow Confident}$ in **Figure S2**), and the probability that an $A^+$ individual adopts the $A^-$ state ($A^+$ to $A^-$ transition probability, given by $A_{\rightarrow Hesitant}$ in **Figure S2**). As with the strength of cultural selection (σ) described previously, the probability that offspring change their vaccine attitude is a function of the $V^+$ frequency in the population. As the frequency of vaccinated individuals ($V^+$) increases in the population, vaccine-confident individuals ($A^+$) are more likely to become hesitant ($A_{\rightarrow Hesitant}$ probability increases) and vaccine-hesitant individuals ($A^-$) are less likely to become confident (

$A_{\rightarrow Confident}$ probability decreases). Similarly to the cultural selection function, the belief transition functions were generated by first choosing a function with a shape that aligned with our general assumptions and then modifying the function to fit specific criteria: 1) probabilities could approach zero, but not equal zero, 2) transition to supporting belief and transition to opposing belief are equally likely at 50% vaccination frequency, and, 3) that high vaccination frequencies (e.g. above herd-immunity levels of vaccination coverage) promote the transition to vaccine hesitancy [5,6]. The upper bound for the belief transition functions were set by calculating the percent difference between vaccine refusal rates in 1991 and 2004 in the United States to estimate transition probabilities between 1–2% [3]. By modulating the attitude transition probabilities according to the vaccination coverage in this manner, we assume that when vaccine coverage ($V^+$ frequency, $x_1 + x_2$) is low, disease occurrence is high and the negative effects of the disease are experienced widely, thus the benefits of being vaccinated (and the costs of not being vaccinated) are more evident [7,8]. As vaccination coverage ($V^+$) increases in the population, and thus disease occurrence is low, the benefits to being vaccinated are less obvious, while low-probability costs such as adverse reactions become more apparent and could be perceived as being riskier than the disease itself. Modulating both the attitude transition probabilities and the cultural selection coefficient according to the level of vaccination coverage in a population reflects that perceptions about the vaccine and its associated effects on health could be meaningfully different in a population with high vaccination coverage than in one with low coverage.

To compute the frequency of a given phenotype in the next iteration, we sum the probability that each mating pair produces offspring of that phenotype over each of the sixteen possible mating pairs. Cultural selection ($\sigma$), described above, then operates on offspring with the $V^+$ trait. At the end of each timestep, the frequency of each phenotype is divided by the sum of all four frequencies, ensuring that the frequencies sum to 1. The full recursions, giving $x_i'$ phenotype frequencies in the next iteration in terms of $x_i$ in the current iteration, are given in **Text S2**. If $x_i'$ is equal to $x_i$, the system is at equilibrium. Unless otherwise stated, the model is initialized with phenotypic frequencies structured to represent those of the United States: $x_1$ (frequency of $V^+A^+$) = 0.81, $x_2$ ($V^+A^-$) = 0.1, $x_3$ ($V^-A^+$) = 0.07, $x_4$ ($V^-A^-$) = 0.02. These frequencies were estimated using reports of Measles-Mumps-Rubella (MMR) vaccination rates and estimates of vaccine attitude frequencies obtained from various sources in the literature [6,9] and the Centers of Disease Control ChildVax database [10,11].

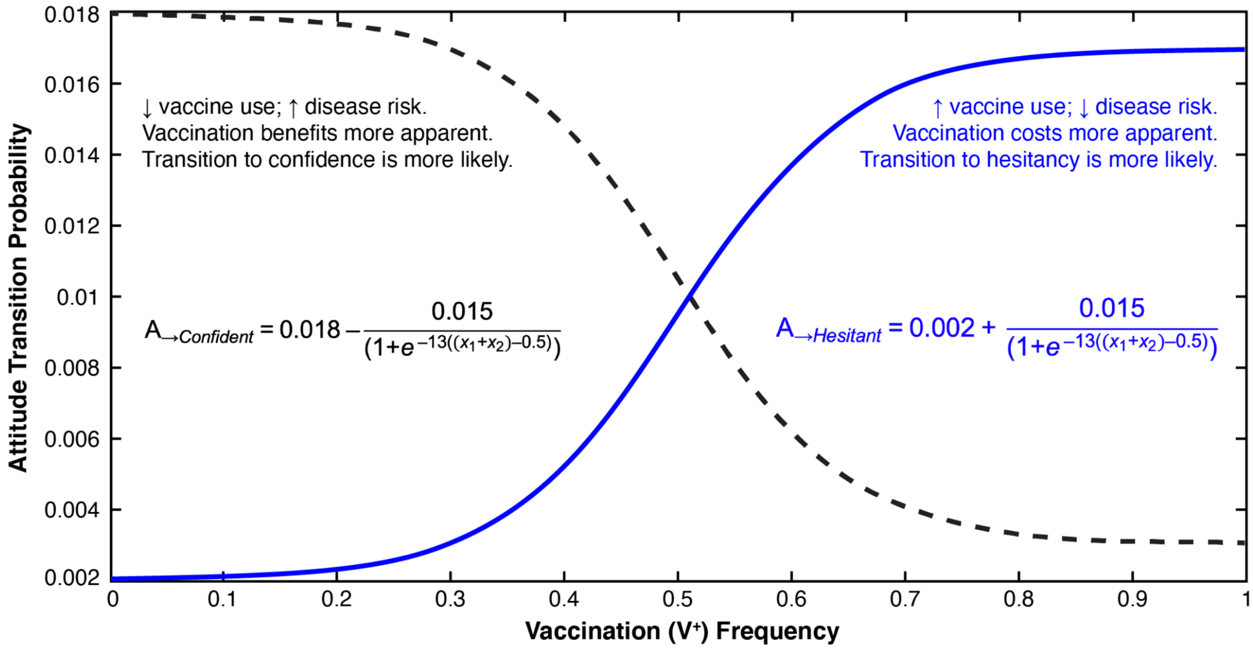


**Figure S2: Attitude Transition Probability Function.** Attitude transition probability functions were constructed by fitting a curve to specified values. Attitude transition probability (vertical axis) is a function of the vaccination frequency in the population ($V^+$; horizontal axis). The probability that a vaccine hesitant individual adopts vaccine confidence ($A^-$ to $A^+$ transition probability, shown in black) is determined by the function $A_{\rightarrow Confident}$, and the probability that a vaccine confident individual adopts vaccine hesitancy ($A^+$ to $A^-$ transition probability, shown with a blue dashed line) is determined by the function $A_{\rightarrow Hesitant}$.

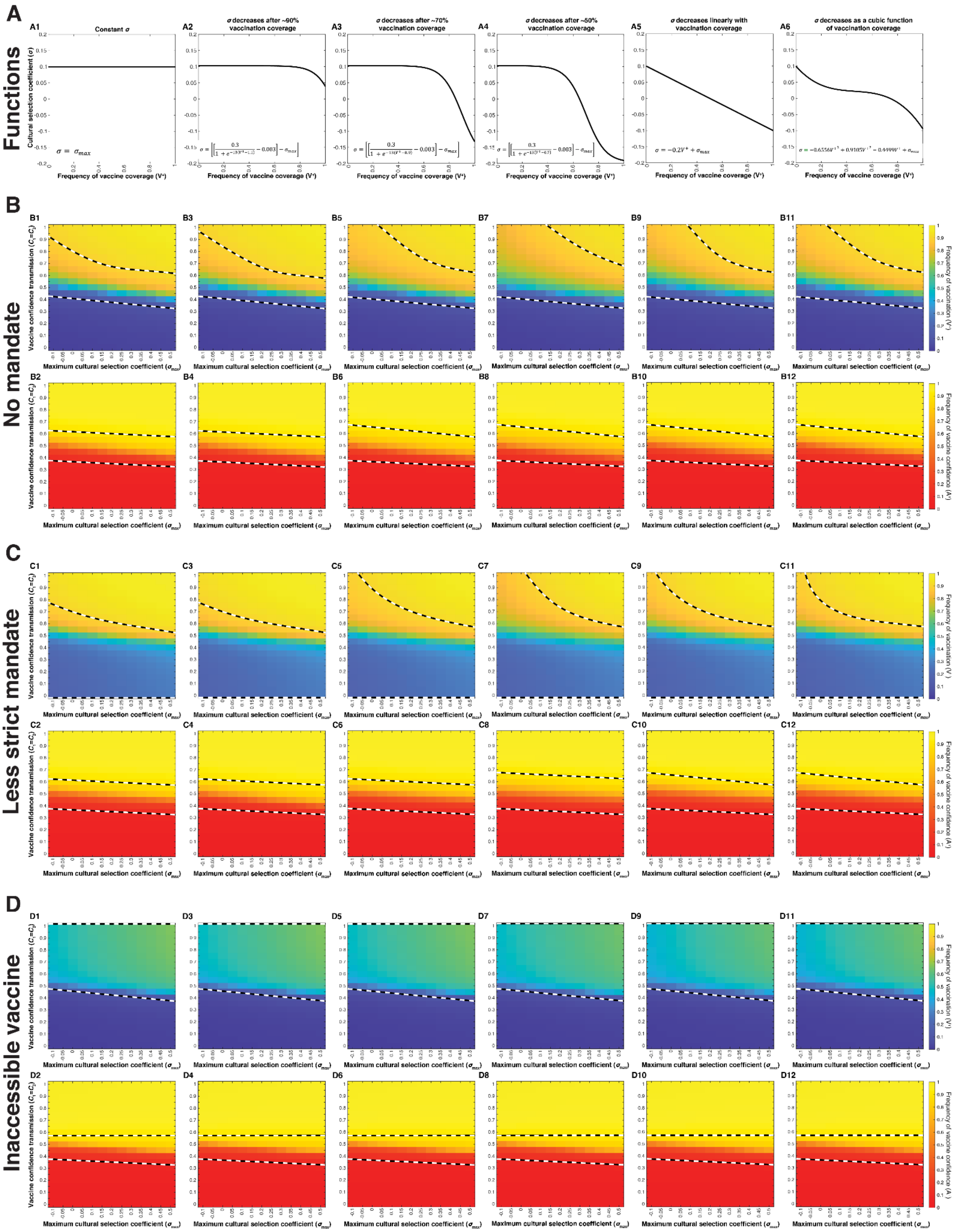

A
Functions
A1 Constant σ
A2 σ decreases after ~90% vaccination coverage
A3 σ decreases after ~70% vaccination coverage
A4 σ decreases after ~50% vaccination coverage
A5 σ decreases linearly with vaccination coverage
A6 σ decreases as a cubic function of vaccination coverage
Cultural selection coefficient (σ)
Frequency of vaccine coverage (V⁺)
σ = σmax
σ = −0.2V⁺ + σmax
B
No mandate
C
Less strict mandate
D
Inaccessible vaccine
Vaccine confidence transmission (C1=C2)
Maximum cultural selection coefficient (σmax)
Frequency of vaccination (V⁺)
Frequency of vaccine confidence (A⁺)

**Figure S3: Selection trajectories affect outcomes at equilibrium when vaccines are accessible..** Heatmaps showing equilibrium vaccine coverage and vaccine confidence levels with an accessible vaccine and no mandate (**Section B**), with an accessible vaccine and a less strict mandate (**Section C**) and an environment with vaccines somewhat inaccessible (**Section D**), employing various cultural selection (σ) functions: (**A1**) σ does not depend on vaccination coverage, (**A2**) σ decreases after a high herd-immunity threshold of ~90% coverage, (**A3**) σ decreases after a medium herd-immunity threshold of ~70% coverage (baseline function), (**A4**) σ decreases after a low herd-immunity threshold of ~50% coverage, (**A5**) σ decreases linearly as vaccination coverage increases, (**A6**) σ decreases according to a cubic function. We vary $C_1 = C_2$ (confidence transmission probability of mixed-attitude couples) on the vertical axis, and maximum selection coefficient $\sigma_{max}$ (indicative of the perceived value of vaccinating offspring) on the horizontal axis. Unspecified parameters are given in **Table 1** with $\sigma_{max}$ held at 0.1 for all functions shown in **Section A** but varied in the heatmaps in **Sections B-D**. Black and white dashed lines indicate the area of the heat maps in which vaccination and confidence frequencies equilibrate between 0.1 and 0.9.

**Table S2: Probabilities of trait transmission to offspring from cultural trait pairings.**
For each mating, we give the probability of transmitting each trait, and corresponding influence parameters. The probability of vaccinating an offspring, $B_{m,n}$, depends on both the parents' vaccination state ($V^+$: vaccinated; $V^-$: unvaccinated) and their attitude state ($A^+$: vaccine confident; $A^-$: vaccine hesitant). $B_{m,n}$ is informed by the influence of parents' vaccination states (**V**) on their decision to vaccinate ($b_m$) and by the influence of their vaccine attitudes (**A**) on their decision to vaccinate ($c_n$). For each parental pairing, the probability of not vaccinating an offspring is $1 - B_{m,n}$. Each pairing transmits confidence in vaccines at a rate $C_n$, and hesitancy at rate $1 - C_n$. The parameters $b_m$, $c_n$, and $C_n$ are set as constants for each simulation, and $B_{m,n}$ is calculated from these.

| | **Trait Transmission Probabilities** | | | | **Influence of parental vaccination and attitudes on offspring vaccination** | |
|---|---|---|---|---|---|---|
| **Mating pair** | **Offspring vaccination ($V^+$) probability** | **$V^-$ offspring probability** | **$A^+$ offspring probability** | **$A^-$ offspring probability** | **V influence (*m*)** | **A influence (*n*)** |
| $V^+A^+ \times V^+A^+$ | $B_{m=3,n=3} = c_3\left(\frac{1+b_3}{2}\right)$ | $1 - B_{3,3}$ | $C_3$ | $1 - C_3$ | $b_3$ | $c_3$ |
| $V^+A^+ \times V^+A^-$ | $B_{3,2} = c_2\left(\frac{1+b_3}{2}\right)$ | $1 - B_{3,2}$ | $C_2$ | $1 - C_2$ | $b_3$ | $c_2$ |
| $V^+A^- \times V^+A^+$ | $B_{3,1} = c_1\left(\frac{1+b_3}{2}\right)$ | $1 - B_{3,1}$ | $C_1$ | $1 - C_1$ | $b_3$ | $c_1$ |
| $V^+A^- \times V^+A^-$ | $B_{3,0} = c_0\left(\frac{1+b_3}{2}\right)$ | $1 - B_{3,0}$ | $C_0$ | $1 - C_0$ | $b_3$ | $c_0$ |
| $V^+A^+ \times V^-A^+$ | $B_{2,3} = c_3\left(\frac{1+b_2}{2}\right)$ | $1 - B_{2,3}$ | $C_3$ | $1 - C_3$ | $b_2$ | $c_3$ |
| $V^+A^+ \times V^-A^-$ | $B_{2,2} = c_2\left(\frac{1+b_2}{2}\right)$ | $1 - B_{2,2}$ | $C_2$ | $1 - C_2$ | $b_2$ | $c_2$ |
| $V^+A^- \times V^-A^+$ | $B_{2,1} = c_1\left(\frac{1+b_2}{2}\right)$ | $1 - B_{2,1}$ | $C_1$ | $1 - C_1$ | $b_2$ | $c_1$ |
| $V^+A^- \times V^-A^-$ | $B_{2,0} = c_0\left(\frac{1+b_2}{2}\right)$ | $1 - B_{2,0}$ | $C_0$ | $1 - C_0$ | $b_2$ | $c_0$ |
| $V^-A^+ \times V^+A^+$ | $B_{1,3} = c_3\left(\frac{1+b_1}{2}\right)$ | $1 - B_{1,3}$ | $C_3$ | $1 - C_3$ | $b_1$ | $c_3$ |
| $V^-A^+ \times V^+A^-$ | $B_{1,2} = c_2\left(\frac{1+b_1}{2}\right)$ | $1 - B_{1,2}$ | $C_2$ | $1 - C_2$ | $b_1$ | $c_2$ |
| $V^-A^- \times V^+A^+$ | $B_{1,1} = c_1\left(\frac{1+b_1}{2}\right)$ | $1 - B_{1,1}$ | $C_1$ | $1\text{-}C_1$ | $b_1$ | $c_1$ |
| $V^-A^- \times V^+A^-$ | $B_{1,0} = c_0\left(\frac{1+b_1}{2}\right)$ | $1 - B_{1,0}$ | $C_0$ | $1\text{-}C_0$ | $b_1$ | $c_0$ |
| $V^-A^+ \times V^-A^+$ | $B_{0,3} = c_3\left(\frac{1+b_0}{2}\right)$ | $1 - B_{0,3}$ | $C_3$ | $1\text{-}C_3$ | $b_0$ | $c_3$ |
| $V^-A^+ \times V^-A^-$ | $B_{0,2} = c_2\left(\frac{1+b_0}{2}\right)$ | $1 - B_{0,2}$ | $C_2$ | $1\text{-}C_2$ | $b_0$ | $c_2$ |
| $V^-A^- \times V^-A^+$ | $B_{0,1} = c_1\left(\frac{1+b_0}{2}\right)$ | $1 - B_{0,1}$ | $C_1$ | $1\text{-}C_1$ | $b_0$ | $c_1$ |
| $V^-A^- \times V^-A^-$ | $B_{0,0} = c_0\left(\frac{1+b_0}{2}\right)$ | $1 - B_{0,0}$ | $C_0$ | $1\text{-}C_0$ | $b_0$ | $c_0$ |

**Table S3: Probability range shift assignments**

Each probability was grouped according to baseline vaccination probability calculations. All probabilities in a group hold the value assigned to that group in the range, as shown. $C_n$ probabilities were assigned values as shown, with $C_0$ taking the lowest value in the range and $C_3$ taking the highest. The lowest probability range group is given as an example of value assignment.

| | Range: Low → High | | | |
|---|---|---|---|---|
| Parameters | $C_0$ | $C_1$ | $C_2$ | $C_3$ |
| Example value (range 0.1–0.4) | 0.1 | 0.2 | 0.3 | 0.4 |

**Table S4: Quantitative differences between equilibrium frequencies with low transmission of vaccine confidence**

The mean and median of vaccination coverage and vaccine confidence levels at equilibrium were calculated for the section of the heatmaps in **Figure 2** for which $C_1 = C_2 < 0.5$ (blue in vaccination coverage heatmaps; red in the confidence level heatmaps).

| Vaccination Coverage below $C_1 = C_2 < 0.5$ | **No Mandate** | **Less Strict Mandate** | **Vaccine Inaccessible** |
|---|---|---|---|
| **Mean** | 9.031% | 27.723% | 5.032% |
| **Median** | 4.047% | 25.872% | 2.578% |
| Confidence Levels below $C_1 = C_2 < 0.5$ | | | |
| **Mean** | 10.875% | 9.092% | 9.927% |
| **Median** | 5.262% | 5.115% | 5.178% |

**Text S2: Recursions for Vaccine Niche Construction**

$$\bar{w}x_1' = (1+\sigma_1)\big(m_{11}B_{3,3}C_3 + m_{12}B_{3,2}C_2 + m_{21}B_{3,1}C_1 + m_{13}B_{2,3}C_3 + m_{31}B_{1,3}C_3 + m_{14}B_{2,2}C_2 + m_{41}B_{1,1}C_1 + m_{22}B_{3,0}C_0 + m_{23}B_{2,1}C_1 + m_{32}B_{1,2}C_2 + m_{24}B_{2,0}C_0 + m_{42}B_{1,0}C_0 + m_{33}B_{0,3}C_3 + m_{34}B_{0,2}C_2 + m_{43}B_{0,1}C_1 + m_{44}B_{0,0}C_0\big)$$

$$\bar{w}x_2' = (1+\sigma_1)\big(m_{11}B_{3,3}(1-C_3) + m_{12}B_{3,2}(1-C_2) + m_{21}B_{3,1}(1-C_1) + m_{13}B_{2,3}(1-C_3) + m_{31}B_{1,3}(1-C_3) + m_{14}B_{2,2}(1-C_2) + m_{41}B_{1,1}(1-C_1) + m_{22}B_{3,0}(1-C_0) + m_{23}B_{2,1}(1-C_1) + m_{32}B_{1,2}(1-C_2) + m_{24}B_{2,0}(1-C_0) + m_{42}B_{1,0}(1-C_0) + m_{33}B_{0,3}(1-C_3) + m_{34}B_{0,2}(1-C_2) + m_{43}B_{0,1}(1-C_1) + m_{44}B_{0,0}(1-C_0)\big)$$

$$\bar{w}x_3' = \big(m_{11}(1-B_{3,3})C_3 + m_{12}(1-B_{3,2})C_2 + m_{21}(1-B_{3,1})C_1 + m_{13}(1-B_{2,3})C_3 + m_{31}(1-B_{1,3})C_3 + m_{14}(1-B_{2,2})C_2 + m_{41}(1-B_{1,1})C_1 + m_{22}(1-B_{3,0})C_0 + m_{23}(1-B_{2,1})C_1 + m_{32}(1-B_{1,2})C_2 + m_{24}(1-B_{2,0})C_0 + m_{42}(1-B_{1,0})C_0 + m_{33}(1-B_{0,3})C_3 + m_{34}(1-B_{0,2})C_2 + m_{43}(1-B_{0,1})C_1 + m_{44}(1-B_{0,0})C_0\big)$$

$$\bar{w}x_4' = \big(m_{11}(1-B_{3,3})(1-C_3) + m_{12}(1-B_{3,2})(1-C_2) + m_{21}(1-B_{3,1})(1-C_1) + m_{13}(1-B_{2,3})(1-C_3) + m_{31}(1-B_{1,3})(1-C_3) + m_{14}(1-B_{2,2})(1-C_2) + m_{41}(1-B_{1,1})(1-C_1) + m_{22}(1-B_{3,0})(1-C_0) + m_{23}(1-B_{2,1})(1-C_1) + m_{32}(1-B_{1,2})(1-C_2) + m_{24}(1-B_{2,0})(1-C_0) + m_{42}(1-B_{1,0})(1-C_0) + m_{33}(1-B_{0,3})(1-C_3) + m_{34}(1-B_{0,2})(1-C_2) + m_{43}(1-B_{0,1})(1-C_1) + m_{44}(1-B_{0,0})(1-C_0)\big)$$
